# INHIBITORY EFFECT OF STRAINS WITH PROBIOTIC POTENTIAL ON PATHOGENIC BACTERIA CAUSING ACUTE DIARRHEA DISEASES

**DOI:** 10.1101/2024.11.18.24317495

**Authors:** Margarita Maria Diaz Durango, Linda Maria Chams Chams, Shaenne Sophia Julio Alvarez, Yuris Esther Ortega Diaz, Jorge Luis Negrete Penata, Leidy Juliet Rios Marin, Rafael G. Villarreal J, Gabriel A. Pérez, Jonny Andrés Yepes, Carlos J. Castro-Cavadia

## Abstract

Lactic acid bacteria are known as agents that help prevent digestive disorders such as acute diarrheal disease, which is one of the main causes of mortality and morbidity in children worldwide, caused mainly by bacteria, viruses and parasites.

**objective:** To evaluate in vitro the inhibitory effect of lactic acid bacteria with probiotic potential isolated from coastal cheese and raw milk on pathogenic bacteria causing acute diarrheal disease. Materials and methods. A cross-sectional descriptive study was carried out. From 78 samples, 93 isolates were made in Man Rogosa Sharpe agar at 37oC for 48 hours in an anaerobic atmosphere. The probiotic capacity was evaluated by tolerance tests to: pH (2, 3, 4, 5 and 6), bile salts (0.05, 0.1, 0.15, 0.3%), and NaCl (2, 4, 7, 10%); growth was determined at temperatures of (30, 37 and 40°C) and the inhibitory effect in vitro against Salmonella enterica subsp enterica ATCC 14028, Staphylococcus aureus ATCC 29213, Escherichia coli ATCC 25922, Yersinia enterocolitica ATCC 23715, Shigella flexneri ATCC 12022 and Listeria monocytogenes ATCC 19115. Strains that met the requirements to be considered possible probiotics were identified biochemically by the standardized API 50 CHL system. Results. Of all the strains evaluated, four showed probiotic potential. With respect to its inhibitory effect, the tests showed inhibition against one or several microorganisms except L. monocytogenes. These strains were identified as Lactobacillus plantarum and Lactobacillus delbrueckii spp delbrueckii. Conclusions. Raw milk and coastal cheese are an important source of isolation for Lactobacillus spp. with probiotic potential, determining a positive effect against inhibition of gastrointestinal pathogens, being able to contribute to the development of strategies for the prevention of acute diarrheal diseases.

**SUMMARY:** Lactic acid bacteria are known as agents that help prevent digestive disorders such as acute diarrheal disease, which is one of the main causes of mortality and morbidity in children worldwide, caused mainly by bacteria, viruses and parasites. Objective. To evaluate in vitro the inhibitory effect of lactic acid bacteria with probiotic potential isolated from coastal cheese and raw milk on pathogenic bacteria causing acute diarrheal disease. Materials and methods. A descriptive cross-sectional study was conducted. From 78 samples, 93 strains were isolated on Man Rogosa Sharpe agar at 37oC for 48 hours in an anaerobic atmosphere. The probiotic capacity was evaluated by tolerance tests to: pH (2, 3, 4, 5 and 6), bile salts (0.05, 0.1, 0.15, 0.3%), and NaCl (2, 4, 7, 10%); its growth was determined at temperatures of (30, 37 and 40 °C) and the in vitro inhibitory effect against *Salmonella enterica*subsp enterica ATCC 14028, *Staphylococcus aureus* ATCC 29213, Escherichia coli ATCC 25922, *Yersinia enterocolitica* ATCC 23715, *Shigella flexneri* ATCC 12022 and *Listeria monocytogenes* ATCC 19115.

Strains that met the requirements to be considered possible probiotics were biochemically identified.using the standardized API system®50 CHL. Results. Of all the strains evaluated, four showed probiotic potential. Regarding their inhibitory effect, the tests showed inhibition against one or more microorganisms except L. monocytogenes. These strains were identified as Lactobacillus plantarum and Lactobacillus delbrueckii spp. delbrueckii. Conclusions. Raw milk and coastal cheese are an important source of isolation of Lactobacillus spp. with probiotic potential, determining a positive effect against the inhibition of gastrointestinal pathogens, and may contribute to the development of strategies for the prevention of acute diarrheal diseases.

## INTRODUCTION

The World Health Organization (WHO) describes Acute Diarrheal Diseases (EDAs)such as passing loose or liquid stools three or more times a day that occur more frequently than usual. The infection is spread through contaminated food or drinking water, or from one person to another as a result of poor hygiene. They are generally infectious and self-limiting in nature, caused by various bacterial and viral organisms and, to a lesser extent, parasites. The most common etiological agent of acute diarrhea in pediatric patients is Rotavirus; followed bybacteria such as: Shigella flexneri, Shigella dysenteriae, Salmonella enteritidis, Campylobacter jejuni, Vibrio cholerae, enteropathogenic Escherichia coli, Yersinia enterocolitica, Staphylococcus aureus, among others; and parasites such as: Giardia lamblia, Entamoeba histolytica, Blastocystis hominis and Cryptosporidium spp. It should be noted that the relative frequency of each microorganism varies according to the socioeconomic conditions of each population (WHO, 2017; Rivero et al., 2017).

It is estimated that 30% of cases of ADD are associated with bacterial agents. In this order of ideas, Salmonella enteritidis is one of the four main causes of ADD worldwide. Although most cases of salmonellosis are mild, sometimes the disease can be fatal. It is estimated that 93.8 million cases of non-typhi Salmonella occur annually and approximately 150,000 deaths occur worldwide. In Colombia in 2017, 852 isolates were reported to the Public Health Laboratory, with Salmonella enterica serotype enteritidis and Salmonella enterica serotype typhimurium being the most prevalent. On the other hand,*Shigella*spp., is the main cause of dysentery in both children and adults worldwide. In addition, it is responsible for approximately 5-10% of cases of watery diarrhea and 30% of cases of dysentery. It should also be mentioned that the frequency of diarrhea caused by pathogenic E. coli in Colombia is not known. Recent reports in the Colombian Caribbean region and in central Colombia indicate that the frequency is 7.5% of all cases of EDA in children under five years of age. However, diarrhea caused by E. coli is much more frequent in low-income countries where it can be up to 50% of all cases (Rivero et al., 2015; Gómez, 2014).

According to the WHO and the United Nations Children’s Fund (UNICEF), EDAs represent the second leading cause of death in children under five years of age, causing the death of 525,000 children each year.The malnourished or immunosuppressed are at greatest risk for potentially fatal diarrheal diseases. In addition, these leadto long-term consequences, including malnutrition, decreased growth and impaired cognitive development (WHO, 2017).

In this order of ideas, in 2017 in Colombia, according to the National Institute of Health (INS), 128 deaths were reported to the Public Health Surveillance System (SIVIGILA), corresponding to a mortality rate due to EDA in children under five years of age of 2.8 deaths per 100,000 children under five years of age (INS, 2017).

In relation to the above, due to their high prevalence and mortality and morbidity rates, EDAs continue to be a serious public health problem worldwide, especially in developing countries, where lack of access to drinking water, inadequate excrement management, overcrowding, poor hygiene, among others, facilitate the onset of the disease and therefore the probability of death from this cause, affecting all age groups, with children under five years of age being the most vulnerable; In addition, the misuse of antibiotics has led to the emergence and spread of resistant bacterial strains, which make treatments for infectious diseases in humans no longer effective (WHO, 2017).

Antimicrobial resistance is one of the main threats facing modern medicine, making it a public health problem; therefore, the search for other effective alternatives against the disease is evident. A timely and profitable option that is currently gaining acceptance in the food industry is the use of probiotic bacteria..

Currently, to define probiotics, the concept declared by the Food and Agriculture Organization of the United Nations (FAO) and the WHO is adopted: “probiotics are living organisms that when administered in adequate quantities provide or generate beneficial effects on the health of the host” (FAO, 2006).

Probiotics are mainly Lactic Acid Bacteria (LAB) belonging to the genera Lactobacillus spp and Bifidobacterium spp; these are known as agents that prevent digestive disorders and diseases; and their oral administration is effective in controlling pathogenic microorganisms. In fact, for a microorganism to be defined as a probiotic it must meet a series of conditions such as: not being pathogenic, being safe and resistant to culinary procedures, gastric acidity and the effect of bile in the duodenum, being able to colonize and adhere to the intestinal mucosa and produce antimicrobial substances to regulate the intestinal microbiota and inhibit the growth of pathogenic bacteria. In addition, they must be able to positively increase immune functions (Gámez et al., 2015; Esparza et al., 2010).

Considering the epidemiological impact of bacterial etiology EDAs in the child population worldwide, and that in Colombia there are very few studies that have demonstrated the influence of strains with probiotic potential on gastrointestinal pathogenic bacteria, it became necessary to know the inhibitory effect of these isolates from unexplored products such as raw milk and coastal cheese, on gastrointestinal pathogenic bacteria of importance for Public Health; thus facilitating new information on their reliability and functionality in vitro, which will help implement strategies to reduce or prevent EDAs based on the antimicrobial properties of these microorganisms.

## MATERIALS AND METHODS

A descriptive cross-sectional study was carried out, with a total of 78 samples (59 of coastal cheese and 19 of raw milk) determined by the statistical formula for finite populations, taking into account as criteria only the sales outlets of coastal cheese registered in the Municipal Health Secretariat and the raw milk sold in the city of Montería.

Statistical formula for finite populations:

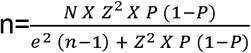

Where:

**N**: Population Size.

**Z:**Confidence level (1.96)

**P**: Expected proportion (50%)

**and:**Allowed error (0.05)

### Isolation of microorganisms

A total of 78 samples were collected, 59 for coastal cheese and 19 for raw milk,which were stored in Ziploc plastic security bags®from SC Johnson and kept in a styrofoam refrigerator under refrigeration and transported within 2 hours to the Food Microbiology Research Laboratory of the Bacteriology Program of the University of Córdoba, for subsequent processing. For the coastal cheese samples, 10 grams were weighed and aseptically macerated; and for the raw milk samples, 10 mL were measured, then homogenized for 5 minutes in 90 mL of a sterile diluent solution (peptone water); subsequently, serial dilutions were prepared in sterile test tubes up to 10-6, keeping the same proportion (9 mL of peptone water and 1 mL of the bacterial suspension). Then, 0.1 mL of the 10-4, 10-5, 10-6 dilutions were inoculated in triplicate in Man Rogosa Sharpe (MRS) culture medium selective for lactic acid bacteria, using the plate extension method and incubated at 37 ° C for 48 hours in an atmosphere with 5% CO2, using envelopes that generate anaerobic conditions. All morphologically different colonies were subcultured and purified using the plate isolation technique in MRS medium and incubated at 37°C for 48 hours under anaerobic conditions. Once isolated, the purity of the strains was verified by microscopic observation of the homogeneity of the cell morphology. For the preliminary selection of the strains, the macroscopic morphology of the colonies and microscopic morphology were taken into account as Gram-positive, non-spore-producing, catalase-negative and non-hemolytic microorganisms (Ramírez et al., 2016; Rodríguez, 2009).

### Evaluation of probiotic potential in vitro

Microorganisms selected according to the morphological characteristics of BAL were evaluated for probiotic potential.performing different in vitro tests in triplicate, according to the methodology cited by Lara and Burgos in 2012 with some modifications (Mantilla et al., 2012).

a. **Tolerance to pH changes**.The viability of the microorganisms was evaluated using MRS broth with different pH values, 2.0, 3.0, 4.0, 5.0 and 6.0 (adjusting the respective pH with HCl or NAOH). The tubes with MRS broth at different pH concentrations were inoculated with a concentration of 0.5 MacFarland scale of the strain under study and incubated at 37°C for 24 hours under anaerobic conditions. Resistance and survival were verified by comparing the viable microorganism count (CFU/mL)(Mantilla et al., 2012).
b. **Tolerance to different concentrations of bile salts**.Tolerance to different concentrations of bile salts was evaluated using MRS broth at 0.05, 0.1, 0.15 and 0.3% w/v of bile salts, adjusting the pH to 6 (pH of the broth), and incubating at 37°C for 24 hours in anaerobiosis. The viability of the strains was determined by counting viable microorganisms (CFU/mL).(Mantilla et al., 2012).
c. **Tolerance to temperature changes**. Tolerance to temperature changes was assessed by incubating the candidate probiotic strains in MRS medium, at temperatures of 30, 37 and 40°C, for 24 hours in an anaerobic jar; The viability of the strains was determined by counting viable microorganisms (CFU/mL).(Mantilla et al., 2012).
d. **Tolerance to different concentrations of NaCl**.The tolerance of the strains under study to different concentrations of NaCl was evaluated using MRS broth with NaCl concentrations of 2, 4, 7 and 10% w/v and incubating them at 37°C for 24 hours; The viability of the strains was determined by counting viable microorganisms (CFU/mL).(Mantilla et al., 2012).
e. **Gas production from glucose**.Gas production was evaluated using 10 mL of MRS broth, with 0.2% v/v of a bromocresol purple solution (0.5%) and Durham hoods; gas production was evidenced by the presence of gas in the hoods, after incubation at 37°C for 24 hours.(Mantilla et al., 2012).

### Inhibitory effect test

The test of the inhibitory effect of strains with probiotic potential against pathogenic bacteria causing EDA was evaluated according to the methodology cited by Agudelo and others with some modifications (Mejía, 2016).

In order to determine the ability of these candidate probiotic strains to inhibit the growth of pathogenic bacteria, pathogenic ATCC strains from the strain bank of the Microbiology Laboratory of the Bacteriology program at the University of Córdoba were used, such as Staphylococcus aureus ATCC 29213, Listeria monocytogenes ATCC 19115, Escherichia coli ATCC 25922, Yersinia enterocolitica ATCC 23715, Salmonella enterica subspecies enterica ATCC 14028, and Shigella flexneri ATCC 12022.

### Activation of ATCC pathogenic strains

Pathogenic strains were reactivated in BHI broth (Brain Heart Infusion)at 37 °C for 24 hours in an aerobic atmosphere. Then each strain was inoculated in their respective solid culture media, as follows: S. aureus and L. monocytogenes in blood agar, E. coli and Y. enterocolitica in MacConkey and S. enterica subsp enterica and S. flexneri in Salmonella-Shigella agar by the surface depletion method. They were then incubated at 37 °C for 24 hours in an aerobic atmosphere. After the incubation time, Gram staining and other tests were performed for each of them, to verify the purity of the strains (Mejía, 2016).

### Conservation and reactivation of strains with probiotic potential

The isolated strains were preserved in MRS broth, supplemented with 30% glycerol and stored at - 80oC in vials for cryopreservation. Before each assay, the strains were reactivated at 37oC for 15 minutes, then 100 were inoculated.μL of the cultures stored in test tubes with 3 mL of MRS broth and incubated for 24 hours at 37 ° C in an anaerobic atmosphere. They were then streaked onto MRS agar at 37 ° C for 48 hours in anaerobiosis. This was done in order to isolate bacterial colonies from each of the cultures and from them perform the cultures necessary to obtain bacterial biomass (Mejía, 2016).

### Preparation of strains with probiotic potential

To prepare the candidate strains for probiotics, filter paper discs (Whatman) were cut®, Grade 3) with a standard perforator and were placed in a clean glass container. Then, it was sterilized in an autoclave at 121°C for 15 minutes. Once the strains were reactivated, the inoculum was adjusted to a concentration of 0.5 on the McFarland scale in MRS broth. Subsequently, with the help of a sterilized clamp, several sterile filter paper discs were taken and added to the suspensions of each strain. Then, they were placed in orbital shaking for 30 minutes and stored at 4oC until use (Ramírez, 2010).

### Preparation of ATCC pathogenic strains

All ATCC pathogenic strains were subcultured on nutrient agar at 37 °C for 24 hours. For each strain, several colonies formed were taken with a sterile loop and placed in a test tube with sterile 0.85% saline solution, which was then homogenized with a vortex mixer. This was repeated until a turbidity similar to that of the McFarland pattern of 0.5 was obtained, which is equivalent to a concentration of approximately 1.5 × 108 CFU/mL. In this way, 6 bacterial suspensions were obtained, one of each pathogenic strain (Mejía, 2016).

To evaluate the inhibitory effect of strains with probiotic potential against various pathogenic bacteria, the Kirby-Bauer method or agar diffusion method was established, carrying out the following procedure: a sterile cotton swab was dipped in the bacterial suspension of each strain of the study (S. aureus, E. Coli, S. enterica subsp enterica, Y. enterocolitica, L. monocytogenes and S. flexneri), and the cotton was pressed against the inner walls of the tube to remove excess liquid. Then, using the same swab, it was massively inoculated on the surface of the Müeller-Hinton agar. Subsequently, after 5-10 minutes, a 6mm diameter filter paper disk impregnated with the liquid culture of the strain with probiotic potential was placed on the dishes inoculated with each of the pathogenic bacteria. Each culture was refrigerated at 4°C for 1 hour and then incubated at 37°C for 24-48 hours in an anaerobic atmosphere. After this time, the presence or absence of growth inhibition halos around the impregnated discs was verified by the formation of transparent zones around the disc. It should be noted that the inhibition zones were measured in millimeters (mm) and the inhibitory effect was calculated by the diameter of the halo measured in mm (Mejía, 2016).

### Biochemical identification of the selected strains

The biochemical identification of the selected strains with probiotic potential was carried out using the standardized API system.®50 CH, composed of 50 microtubes containing different dehydrated substrates intended for the study of carbohydrate metabolism in microorganisms. The API®50 CH is used in combination with the API®50 CHL Medium for the identification of the genus Lactobacillus and other lactic acid bacteria. The procedure was carried out according to the methodology described in the system’s technical data sheet. Initially, several colonies were taken from the culture with the help of a sterile brush and a dense suspension of the microorganism to be studied was made in a tube with 2 mL of sterile distilled water without additives. Then, in another tube with 5 mL of sterile 0.85% saline solution, a suspension of turbidity equal to McFarland pattern 2 was made by transferring with the help of a sterile Pasteur pipette, a certain number of drops of the previous dense suspension and noting said number of drops. Later, in an API ampoule®50 CHL Medium was inoculated twice the number of drops mentioned, homogenized and then immediately inoculated into each tube of the gallery, sealing it with sterile mineral oil to ensure anaerobiosis. Finally, it was incubated for 48 hours at 36ºC +/-2ºC in aerobiosis. During incubation, the catabolism of carbohydrates produces organic acids that turn the pH indicator (bromocresol purple) from purple to yellow contained in the medium and esculin gives a change to black color; the test is considered positive. The first microtube serves as a negative control since it does not contain active ingredient (Ramírez, 2016).

### Statistical analysis

Qualitative variables were analyzed using frequency distribution and quantitative variables were analyzed using measures of central tendency. The Shapiro-Wilk test was applied to assess data normality. The Mann-Whitney U test and Kruskal-Wallis test were also applied to assess nonparametric data. All data were analyzed using GraphPad Prism 8 software.

## RESULTS

From the 78 samples processed between coastal cheese and raw milk, a total of 93 strains were isolated taking into account the morphology of the colonies, showing small and large, circular and punctate colonies, bright white in color, with smooth and wavy edges, convex and with a creamy consistency.

Regarding the microscopic characteristics of the 93 strains isolated in the processed samples, 94.62% (88) were compatible with the morphology of LAB, Gram-positive bacilli, cocci or coccobacilli, while 5.38% (5) corresponded to Gram-negative bacilli. Regarding the Schaeffer-Fulton stain, it was confirmed that 94.62% (88) of the isolates were negative, which is consistent with the characteristics of LAB, since they do not form endospores; while 5.38% (5) were spore-forming.

Regarding the catalase test, 84.94% (80) of the isolated strains were catalase negative, which coincides with the characteristics of the BAL, since the production of bubbles in the presence of hydrogen peroxide was not observed, because they lack the enzyme, while 14.05% (13) were catalase positive. Regarding hemolytic activity, of the total number of isolated strains, 48.38% (45) were found to beγ-hemolytic, because no clear or partially clear area was observed around the colonies on blood agar. These results coincide with the characteristics of LAB. The remaining 51.61% (48) presented hemolysis, which makes them unsuitable for probiotics, since this property is undesirable in the selection criteria because microorganisms that produce hemolysins can use hemoglobin, hemin and hematin from red blood cells as a source of iron (see figure 1). Of the 93 strains isolated, only 25 (27%) met all the characteristics preliminary requirements to be considered as potential probiotics: being Gram positive, catalase negative, non-spore forming and not presenting hemolytic activity (see table 1).

**Table 1.** Criteria for the selection of potential probiotic strains.

| STRAIN | CATALASE | GRAM | FULTON | HEMOLYSIS |
| --- | --- | --- | --- | --- |
| 18 | Negative | Gram Positive Cocci | Negative | Negative |
| 19 | Negative | Gram Positive Cocci | Negative | Negative |
| 20 | Negative | Gram Positive Cocci | Negative | Negative |
| 23 | Negative | Gram Positive Cocci | Negative | Negative |
| 26 | Negative | Gram Positive Cocci | Negative | Negative |
| 33 | Negative | Gram Positive Cocci | Negative | Negative |
| 43G | Negative | Gram Positive Bacilli | Negative | Negative |
| 45 | Negative | Gram-positive coccobacilli | Negative | Negative |
| 50 | Negative | Gram Positive Cocci | Negative | Negative |
| 51P | Negative | Gram Positive Cocci | Negative | Negative |
| 52 | Negative | Gram Positive Cocci | Negative | Negative |
| 53 | Negative | Gram-positive coccobacilli | Negative | Negative |
| 54P | Negative | Gram Positive Cocci | Negative | Negative |
| 55P | Negative | Gram Positive Cocci | Negative | Negative |
| 55G | Negative | Gram Positive Cocci | Negative | Negative |
| 57 | Negative | Gram-positive coccobacilli | Negative | Negative |
| 61 | Negative | Gram Positive Cocci | Negative | Negative |
| 62 | Negative | Gram-positive coccobacilli | Negative | Negative |
| 64 | Negative | Gram Positive Cocci | Negative | Negative |
| 65 | Negative | Gram Positive Cocci | Negative | Negative |
| 66 | Negative | Gram Positive Cocci | Negative | Negative |
| 67 | Negative | Gram Positive Cocci | Negative | Negative |
| 69 | Negative | Gram-positive coccobacilli | Negative | Negative |
| 76P | Negative | Gram Positive Cocci | Negative | Negative |
| 79 | Negative | Gram Positive Cocci | Negative | Negative |

Source: made by the authors

**Figure 1.**
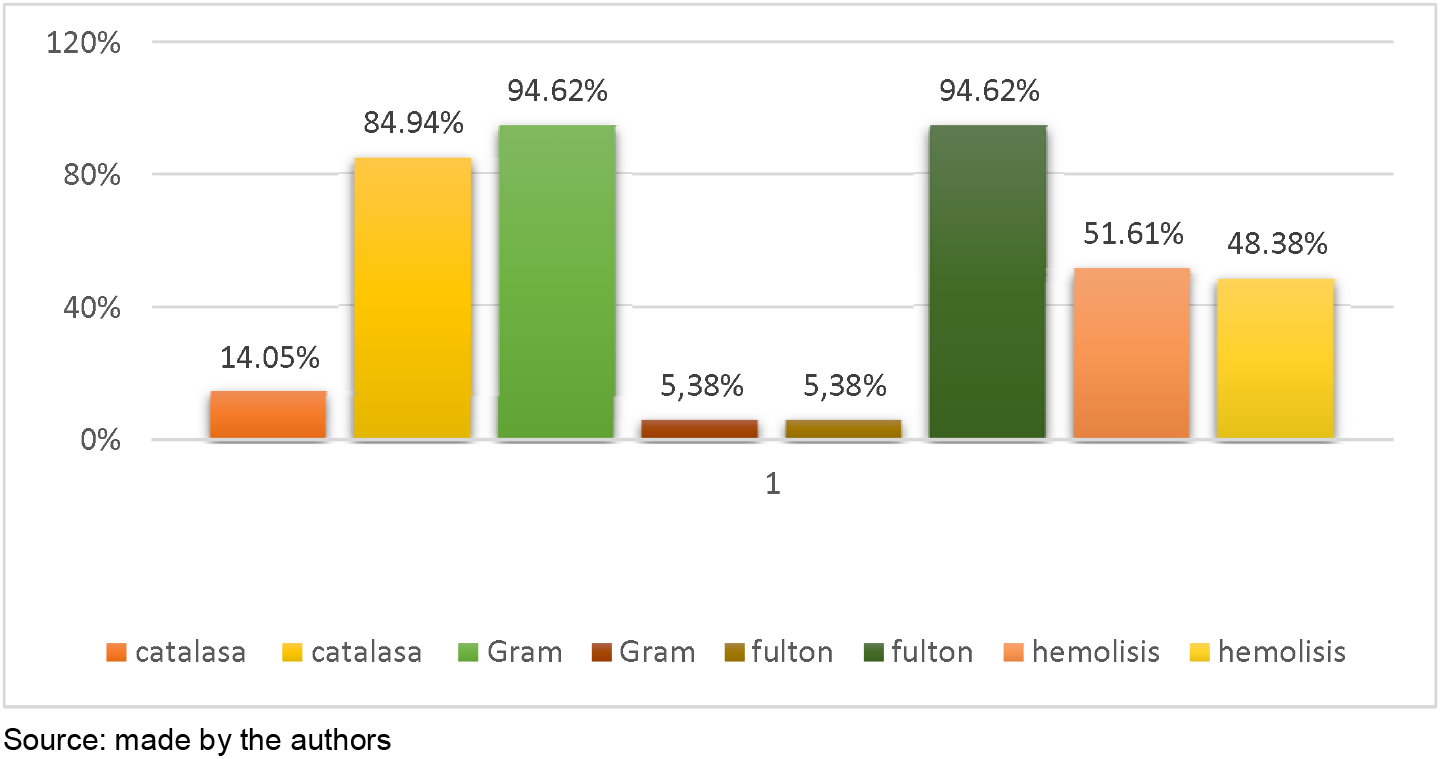
Identification tests and preliminary selection

Regarding the tolerance test at different pH concentrations in the 25 strains selected in the preliminary tests, 4% (6) of them tolerated the MRS broth adjusted to pH 2, 36% (9) to pH 3, 76% (19) to pH 4, 88% (22) pH 5 and 100% (25) pH 6. Significant differences were found (p<0.05)between pH 2 and 3 with pH 5 because there was greater growth at pH 5 (7.2 x108 CFU/mL) compared to pH 2 (872 CFU/mL), and pH 3 (1.2×104 CFU/mL) (See Figure 2). Taking into account the results obtained, it was evident that approximately 30% of the BAL evaluated resisted these low pH concentrations.

**Figure 2.**
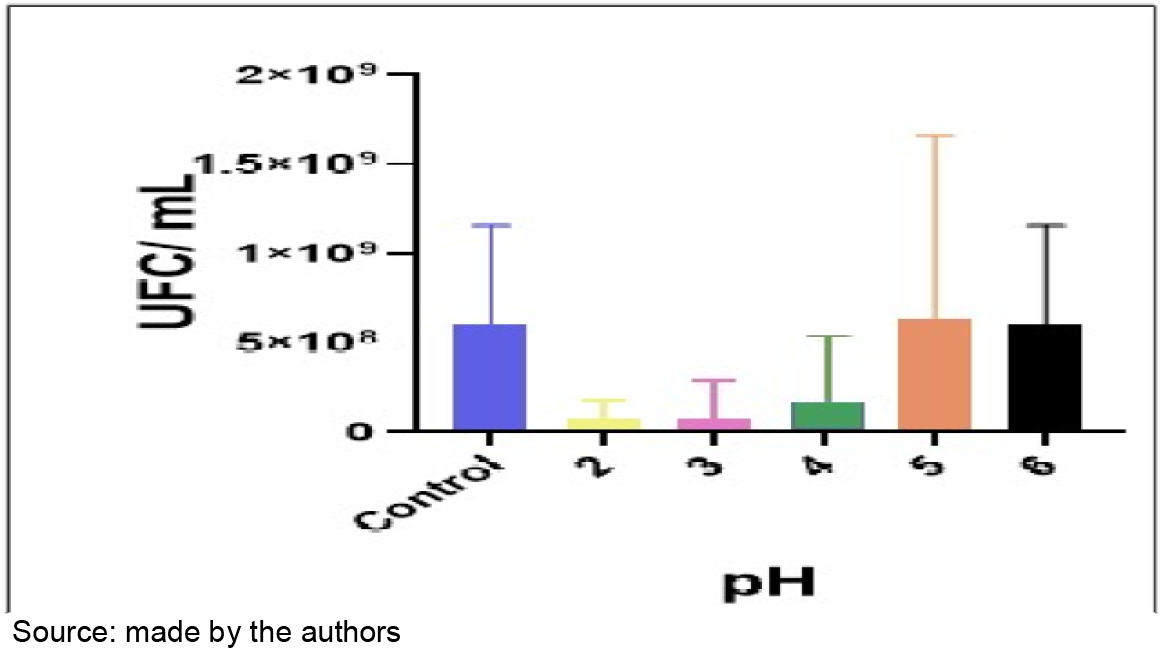
Behavior of LAB at different pH concentrations.

Figure 3 shows that 88% (22/25) of the evaluated BAL tolerated the different concentrations of bile salts to which they were subjected, finding significant differences (p<0.05)between bile salt concentrations of 0.05% (1.2 × 108 CFU/mL) and 0.10% (8.7 × 107 CFU/mL) versus 0.30% (2.7×106 CFU/mL).

**Figure 3.**
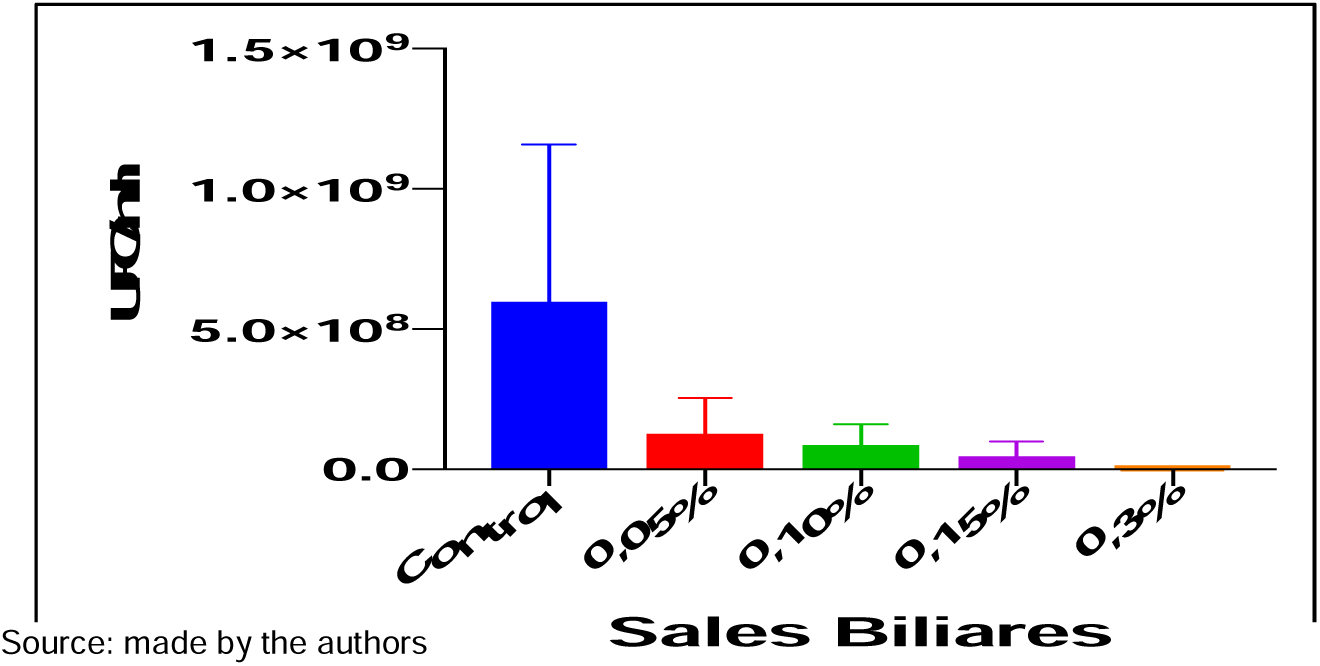
Behavior of BAL at different concentrations of bile salts

Regarding the test of tolerance to different temperatures, 32% (8) of these strains grew at 30°C, 100% (25) at 37°C and 36% (9) at 40°C. Significant differences (p<0.05) were obtained between the temperature of 30°C (8.6 × 106 CFU/mL) and 37°C (8.1 × 107 CFU/mL) as well as between the temperature of 37°C (8.1 × 107 CFU/mL) and 40°C (4.8 × 107 CFU/mL) which shows the effect of temperature on the growth of each microorganism; the temperature of 37°C was the one that showed the best results (see figure 4). Regarding the data obtained, it is worth highlighting the capacity of some strains to tolerate different temperature changes, especially those that resisted 40oC. which allows them to remain viable during processing, storage and distribution.

**Figure 4.**
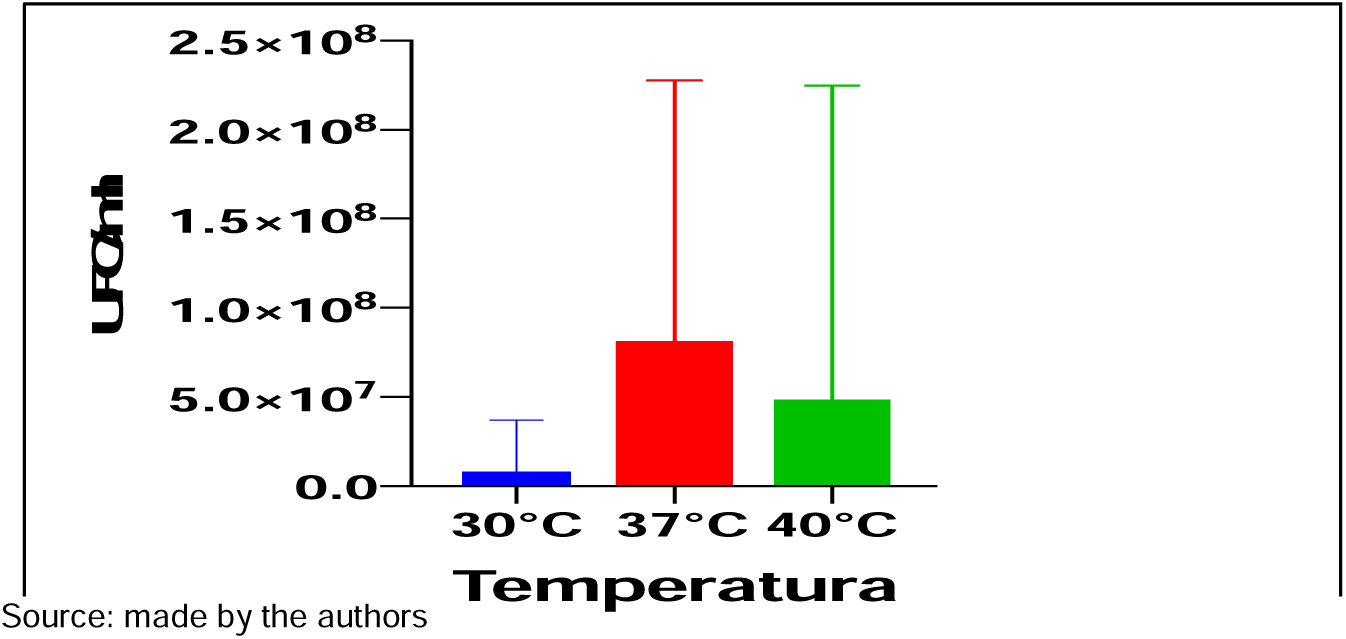
Behavior of BAL at different temperatures.

In the tolerance test to different concentrations of NaCl, 40% (10) of the strains grew in the different concentrations of NaCl evaluated, finding significant differences (p<0.05)between NaCl concentrations of 4% (1.1×108CFU/mL) and 10% (3.7×107 CFU/mL), showing greater tolerance at low concentrations than at high concentrations, however the greatest microbial development occurred at the 7% NaCl concentration as seen in Figure 5.

**Figure 5.**
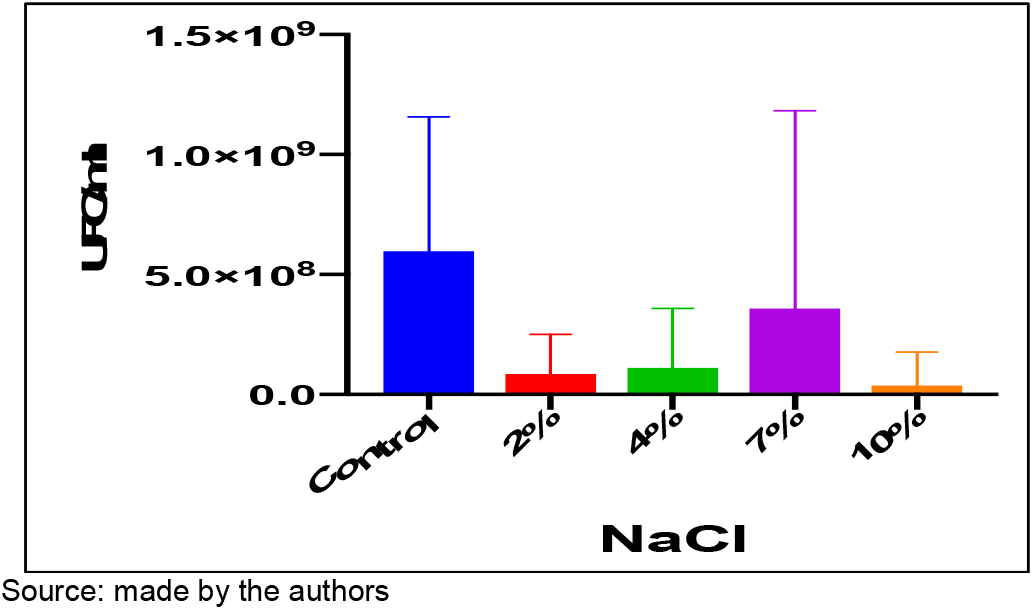
Behavior of BAL at different NaCl concentrations.

Regarding the gas production test from glucose, 100% (25) of the strains evaluated fermented the carbohydrate without gas production, demonstrating its assimilation (See figure 6).

**Figure 6.**
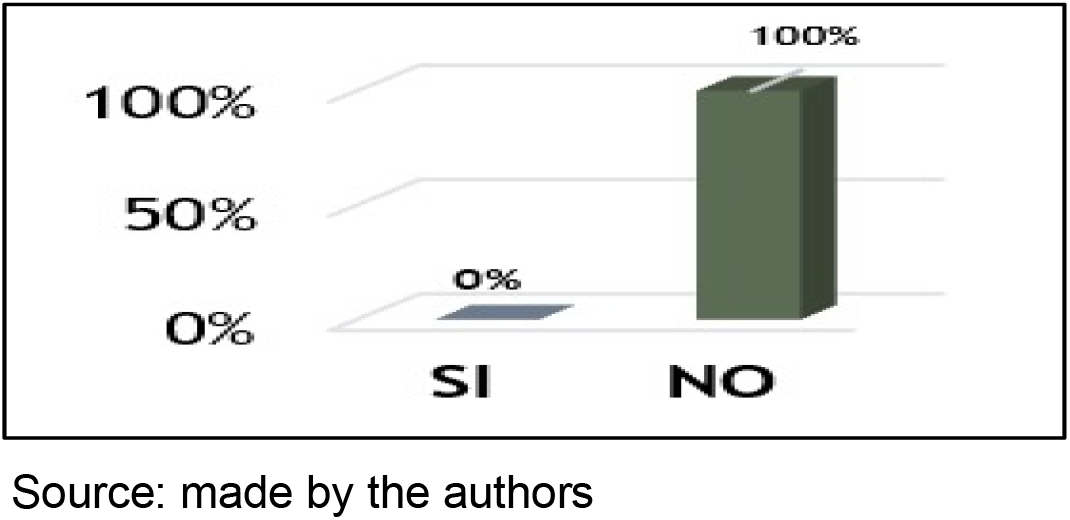
Gas production from glucose

In relation to the test of the inhibitory effect of strains with probiotic potential against pathogenic bacteria, it was shown that 28% (7/25) presented inhibition against Staphylococcus aureus, 20% (5/25) on Shigella flexneri, 16% (4/25) on Yersinia enterocolitica, 12% (3/25) on Escherichia coli, 8% (2/25) on Salmonella enterica subsp enterica and 4% (1/25) on Listeria monocytogenes. It should be noted that 20% (5/25) of the strains presented inhibitory activity against more than one pathogenic bacteria; while 80% (20/25) presented antimicrobial activity against one pathogen. The inhibition halos were variable for each of the microorganisms evaluated. Strains that demonstrated the best antagonistic activity were 18, 52, 62 and 69; and to a lesser extent, strains 45 and 53. Likewise, it was observed that 9 strains (36%) did not present antimicrobial activity against the six pathogens evaluated, while 16 strains (64%) showed activity against one or more pathogens (see Table 2).

**Table 2.** Inhibitory activity against gastrointestinal pathogens.

| #<br>Strain/<br>halo | *Escherichia coli*<br>ATCC 25922 |  | *Salmonella enterica* subsp.<br>*enterica* ATCC 14028 |  | *Listeria monocytogenes*<br>ATCC 19115 |  | *Shigella flexneri*<br>ATCC 12022 |  | *Yersinia enterocolitica*<br>ATCC 23715 |  | *Staphylococcus aureus*<br>ATCC 29213 |  |
| --- | --- | --- | --- | --- | --- | --- | --- | --- | --- | --- | --- | --- |
|  | 24h<br>(mm) | 48h<br>(mm) | 24h<br>(mm) | 48h<br>(mm) | 24h<br>(mm) | 48h<br>(mm) | 24h<br>(mm) | 48h<br>(mm) | 24h<br>(mm) | 48h<br>(mm) | 24h<br>(mm) | 48h<br>(mm) |
| 18 | 0 | 0 | 0 | 0 | 0 | 0 | 8 | 8 | 10 | 10 | 0 | 0 |
| 19 | 0 | 0 | 0 | 0 | 0 | 0 | 0 | 0 | 0 | 0 | 0 | 0 |
| 20 | 0 | 0 | 0 | 0 | 0 | 0 | 0 | 0 | 0 | 0 | 0 | 0 |
| 23 | 0 | 0 | 0 | 0 | 0 | 0 | 0 | 0 | 0 | 0 | 0 | 0 |
| 26 | 0 | 0 | 0 | 0 | 0 | 0 | 0 | 0 | 0 | 0 | 0 | 0 |
| 33 | 0 | 0 | 0 | 0 | 0 | 0 | 0 | 0 | 0 | 0 | 11 | 11 |
| 43G | 0 | 0 | 0 | 0 | 0 | 0 | 0 | 0 | 0 | 0 | 0 | 0 |
| 45 | 0 | 0 | 0 | 0 | 0 | 0 | 0 | 0 | 0 | 0 | 16 | 16 |
| 50 | 0 | 0 | 0 | 0 | 0 | 0 | 0 | 0 | 10 | 10 | 0 | 0 |
| 51P | 0 | 0 | 0 | 0 | 0 | 0 | 0 | 0 | 0 | 0 | 0 | 0 |
| 52 | 0 | 0 | 0 | 0 | 0 | 0 | 8 | 8 | 0 | 0 | 10 | 10 |
| 53 | 0 | 0 | 0 | 0 | 0 | 0 | 0 | 0 | 0 | 0 | 10 | 10 |
| 54 | 0 | 0 | 0 | 0 | 0 | 0 | 0 | 0 | 0 | 0 | 0 | 10 |
| 55P | 0 | 0 | 0 | 0 | 0 | 0 | 0 | 0 | 0 | 0 | 0 | 0 |
| 55G | 0 | 0 | 0 | 0 | 0 | 0 | 8 | 8 | 0 | 0 | 0 | 0 |
| 57 | 0 | 0 | 0 | 0 | 0 | 0 | 0 | 0 | 14 | 14 | 10 | 11 |
| 61 | 0 | 0 | 0 | 0 | 0 | 0 | 0 | 0 | 0 | 0 | 0 | 0 |
| 62 | 8 | 8 | 12 | 12 | 0 | 0 | 0 | 0 | 0 | 0 | 0 | 0 |
| 64 | 11 | 11 | 0 | 0 | 0 | 0 | 0 | 0 | 0 | 0 | 0 | 0 |
| 65 | 0 | 0 | 0 | 0 | 0 | 0 | 0 | 0 | 0 | 0 | 0 | 0 |
| 66 | 0 | 0 | 0 | 0 | 0 | 0 | 0 | 0 | 0 | 0 | 9 | 9 |
| 67 | 11 | 11 | 0 | 0 | 0 | 0 | 0 | 0 | 0 | 0 | 0 | 0 |
| 69 | 0 | 0 | 22 | 22 | 0 | 0 | 8 | 8 | 8 | 8 | 0 | 0 |
| 76P | 0 | 0 | 0 | 0 | 9 | 9 | 0 | 0 | 0 | 0 | 0 | 0 |
| 79 | 0 | 0 | 0 | 0 | 0 | 0 | 10 | 10 | 0 | 0 | 0 | 0 |

Source: made by the authors

Regarding the biochemical identification of the selected BAL, those strains that showed the best performance in terms of resistance to acidic pH between 2 and 3, growth capacity in 0.3% of bile salts, tolerance to high concentrations of NaCl (7% and 10%) and inhibition of gastrointestinal pathogens were taken into account. Considering the above,Four strains (45, 53, 62 and 69) were selected for biochemical identification using the standardized API system.®50 CH. The results obtained after 48 hours of incubation constituted the biochemical profile of the strains, allowing the identification of the microorganism with the help of the apiwebTM identification software. The genera identified were Lactobacillus plantarum in 75% and Lactobacillus delbrueckii spp delbrueckii in 25%, with a reliability percentage of 99.4% and 99.9% respectively (See table 3).

**Table 3.** Identification of LAB with probiotic potential.

| STRAIN | GENUS AND SPECIES | RELATIVE FREQUENCY<br>% |
| --- | --- | --- |
| 45 | *Lactobacillus delbrueckii* spp *delbrueckii* | 25% |
| 53 | *Lactobacillus plantarum* | 25% |
| 62 | *Lactobacillus plantarum* | 25% |
| 69 | *Lactobacillus plantarum* | 25% |

Source: made by the authors

## DISCUSSION

EDAs continue to be a public health problem worldwide,if we consider that it causes around 20% of all deaths that occur in children under five years of age in the world. According to scientific evidence, the most common bacteria causing EDA are E. coli, Salmonella spp, S. flexneri, Y. enterocolitica, among others. Although it is true that oral hydration schemes continue to be considered the cornerstone of treatment, they do not reduce the volume of bowel movements and therefore do not shorten the duration of diarrheal episodes; which leads to high rates of hospitalization, complications, mortality and prolonged stays. For this reason, various therapeutic adjuvants have been established such as probiotics that, when administered in conjunction with rehydration therapy, reduce the duration and severity of acute infectious diarrhea. Many studies have shown the benefits of probiotics for the prevention of EDA.For this reason, the WHO and FAO in 2006, established selection criteria for these microorganisms. One of the criteria and prerequisite for microorganisms to develop their beneficial effects in the intestine is resistance to stomach acidity and bile salts (FAO, 2006; Gutiérrez et al., 2015; Pazos et al., 2013).

LThe BALs identified in this research have the ability to develop in very low acidic pH (2,3,4 and 5), a characteristic of the genus*Lactobacillus*, which is attributed to the presence of a constant gradient between the extracellular pH and the cytoplasmic pH; allowing them to survive the barriers of gastrointestinal transit, including the pH of the stomach, which ranges from approximately 1.5 - 3. These results indicate that if humans ingest probiotic bacteria, they must remain viable after passing through the stomach, therefore their probiotic action will be effective against pathogenic bacteria present in the intestine (León, 2012). The results obtained differ from those of Sánchez et al. (2011) where they evaluated the in vitro probiotic potential of 24 strains of Lactobacillus spp. isolated from the vagina of healthy women, where 16.6% were sensitive to acidic pH concentrations. However, the results are similar to those reported by Gámez et al. (2015) where the viability of Lactobacillus plantarum was determined at different pH (2.5, 4.5 and 7); indicating that the strain is resistant to modifications in the acidity of the medium, obtaining growth at each concentration. Likewise, the LAB evaluated also showed tolerance to different concentrations of bile salts. This is noteworthy, since a strain must not only survive the acidic conditions of the stomach but also bile salts, since the concentrations of these in the small intestine are high (0.2% - 2.0% w/v), variable and difficult to predict, and as their concentration increases, it is important that they can develop their metabolic activities without being completely inhibited, in order to exert beneficial effects in the intestine (Mantilla et al., 2012; León, 2012).

On the other hand, the optimal growth temperature of LAB is 37°C, the temperature at which all the LAB isolated in this research grew. This extrinsic factor is one of the most important that affect the growth and survival of microorganisms, it varies between different genera and reflects the optimal temperature range of their natural habitat; as the temperature rises, the chemical and enzymatic reactions are faster and growth accelerates, to the point where inactivation reactions take place. Indeed, it is important that LAB are able to withstand extreme temperatures and remain viable during processing, storage and distribution of products, so that it exerts beneficial effects on the consumer (Mantilla et al., 2012). These results are similar to those obtained by Sánchez, et al. (2015) who evaluated the in vitro probiotic potential of 17 strains of°Lactobacillus spp.isolated from the intestinal tract of neonatal calves where the optimal growth temperature was 37C.°

Another distinctive feature of LAB is growth in high NaCl concentrations. In this study, the strains showed growth in all NaCl concentrations evaluated, developing better in 7 and 10% NaCl. These results can be related to the source of extraction of the strains, since coastal cheese is characterized by having a high concentration of salt. The results obtained differ from those found by Rondón et al. (2008), where the characterized strains showed good growth in NaCl concentrations of 2 and 4%, very low growth at 7% and no growth was detected in MRS broth at 10% NaCl.

Regarding the test for gas production from glucose, all strains evaluated were negative. This characteristic is important in the evaluation of BAL with probiotic potential, since it indicates that strains that do not produce gas do not alter the functions of the host’s gastrointestinal tract (Gámez et al., 2015).

These results are comparable with those obtained by Rondón et al. (2008), who selected 20 strains of Lactobacillus spp., with probiotic potential, where none of the strains produced gas from the fermentation of glucose.

Probiotic LAB have various effects on the host, one of them is the protection that these microorganisms provide to the digestive tract by producing different antimicrobial substances such as organic acids, diacetyl, acetoin, hydrogen peroxide, reuterin, reuterocycline, antifungal peptides and bacteriocins; which have been used for medical purposes and as biopreservatives in food products in order to inhibit the growth of pathogenic Gram-positive and Gram-negative bacteria, as well as yeasts. This effect is also a desirable characteristic in bacteria for probiotic use, since one of the main therapeutic benefits provided by these microorganisms to the host is the balance of the intestinal microbiota and the control of gastrointestinal infections (Huertas, 2010; Guadarrama et al., 2018).

Indeed, the study of LAB plays an important role in Public Health, since it constitutes an alternative to inactivate pathogens that produce EDAs, through the use of antimicrobial substances, making them capable of inhibiting a wide variety of pathogenic bacteria such as E. coli, Salmonella spp. and Listeria. The inhibitory power of the strains evaluated in the study agree with those reported by Gámez et al. (2015), who determined the inhibitory effect of Lactobacillus plantarum against pathogenic strains of S. aureus, Salmonella typhimurium and E. coli; indicating greater inhibition on S. aureus and less effect on E. coli and S. typhimurium. However, the results differ from those obtained by Bustamante and Alvarado (2015), who evaluated the inhibitory capacity of 7 strains of Lactobacillus spp. isolated from the faeces of newborns against S. aureus, Salmonella enteritidis and Salmonella typhi, showing an inhibitory effect on the growth of S. enteritidis and S. typhi, but not on the growth of S. aureus. This could possibly have been caused by a reduction in pH due to the production of low molecular weight metabolites which can easily penetrate the outer membrane of the Gram-negative bacteria or by various mechanisms such as competition for nutrients and competitive exclusion.

On the other hand, the taxonomic study determined that the four selected strains belong to Lactobacillus plantarum and Lactobacillus delbrueckii spp. delbrueckii; thus, the hypothesis that there are lactic acid bacteria with probiotic potential in coastal cheese and raw milk is confirmed, providing the basis for further research in the field of the use of probiotics as a potential alternative with a high level of specificity both in preventive and adjuvant treatments for infections caused by pathogenic bacteria of importance for Public Health. These results are comparable with the studies carried out by Ramírez and Vélez (2016) who isolated and characterized 18 autochthonous strains from artisanal fresh cheese and raw goat milk, where 38.88% (7 strains) of them were *Lactobacillus plantarum*.However, the results obtained differ from those reported by Cueto et al. (2010), who evaluated a group of 53 lactic acid bacteria isolated from coastal serum, identifying 7 strains as*Enterococcus faecium,Enterococcus durans,Lactobacillus rhamnosus,andLactobacillus fermentum*.

Furthermore, the fact that the strains are producers of antimicrobial substances determines a positive effect against gastroenteritis caused by strains of E. coli, Salmonella spp, S. flexneri and Y. enterocolitica, and can contribute to the reduction of these in a considerable way. This makes the antimicrobial substance produced by L. plantarum and L. delbrueckii subsp. delbrueckii have a promising application as it is active against some of these pathogens.

## CONCLUSIONS

Four strains were selected as potential probiotics due to their performance in in vitro probiotic tests. The taxonomic study determined that strains 53, 62 and 69 belong to Lactobacillus plantarum, showing an inhibitory effect in vitro on the growth of S. aureus, Y. enterocolitica, S. flexneri, Salmonella enterica subsp. enterica and E. coli; and strain 45 corresponds to Lactobacillus delbrueckii subsp. delbrueckii, showing inhibitory activity on the growth of S. aureus. It should be noted that none of the strains showed an inhibitory effect on L. monocytogenes.

Raw milk and coastal cheese constitute an important source of isolation of Lactobacillus with probiotic potential.

The results obtained in this study can be used as a reference for future research on the use of probiotics; they may also be useful in the development of control strategies against acute diarrheal diseases.

## Data Availability

All data produced in the present study are available upon reasonable request to the authors

## ACKNOWLEDGEMENTS

We would like to thank Dr. Linda Chams, Jorge Arrieta and William Hoyos for their support, dedication and advice during the project; and the Microbiological and Biomedical Research Group of the University of Córdoba (GIMBIC) for funding the research.

